# Activities of Daily Living with Bionic Arm Improved by Combination Training and Latching Filter in Prosthesis Control Comparison

**DOI:** 10.1101/2020.10.21.20217026

**Authors:** Michael D. Paskett, Mark R. Brinton, Taylor C. Hansen, Jacob A. George, Tyler S. Davis, Christopher C. Duncan, Gregory A. Clark

## Abstract

**Background:** Advanced prostheses can restore function and improve quality of life for individuals with amputations. Unfortunately, most commercial control strategies do not utilize the rich control information from residual nerves and musculature. Continuous decoders can provide more intuitive prosthesis control from multi-channel neural or electromyographic recordings. Three components influence continuous decoder performance: the data used to train the algorithm, the algorithm, and smoothing filters on the algorithm’s output. As individual groups often focus on a single decoder, very few studies compare different decoders using otherwise similar experimental conditions.

**Methods:** We completed a two-phase head-to-head comparison of 12 continuous decoders using activities of daily living. In phase one, we compared two training types and a smoothing filter with three algorithms (modified Kalman filter, multi-layer perceptron, and convolutional neural network) in a clothespin relocation task. We compared training types that included data with only individual digit and wrist movements vs. combination movements (e.g., simultaneous grasp and wrist flexion). We also compared raw vs. nonlinearly smoothed algorithm outputs. In phase two, we compared the three algorithms in fragile egg, zipping, pouring, and folding tasks using the combination training and smoothing found beneficial in phase one. In both phases, we collected objective, performance-based (e.g., success rate) and subjective, user-focused (e.g., preference) measures.

**Results:** Phase one showed that combination training improved prosthesis control accuracy and speed, and that the nonlinear smoothing improved accuracy but generally reduced speed. Phase one importantly showed simultaneous movements were used in the task, and that the modified Kalman filter and multi-layer perceptron predicted more simultaneous movements than the convolutional neural network. In phase two, user-focused metrics favored the convolutional neural network and modified Kalman filter, whereas performance-based metrics were generally similar among all algorithms.

**Conclusions:** These results confirm that state-of-the-art algorithms, whether linear or nonlinear in nature, functionally benefit from training on more complex data and from output smoothing. These studies will be used to select a decoder for a long-term take-home trial with implanted neuromyoelectric devices. Overall, clinical considerations may favor the mKF as it is similar in performance, faster to train, and computationally less expensive than neural networks.

## Background

Modern prosthetic hands can now recreate the complex movements of the human hand [1]–[8]. Unfortunately, unintuitive control makes prostheses difficult to use [9] and is a major factor in prosthesis dissatisfaction and abandonment [10]. Neural and electromyographic (EMG) signals from residual muscles provide a rich source of movement information that could be used to provide intuitive prosthesis control. With rare exceptions [11], [12], commercial prosthetic control uses only two-electrode EMG setups that provide sequential control of up to two degrees-of-freedom [1], [3], [5], [6], [13], [14], far inferior to the abilities of modern prosthetic hands.

Using more neuromuscular inputs can provide more intuitive prosthesis control through richer, more diverse data that can be classified into different movements based on residual neuromuscular activation patterns. Several groups have demonstrated intuitive prosthesis control with decoders that classify distinct movements [15]–[22]. Generally, these decoders classify EMG patterns to several pre-determined grip patterns (e.g., close hand, open hand, pinch). More advanced classification strategies incorporate proportionality into the classes (e.g., enable partial hand closure) [23] or allow simultaneously active classes (e.g., simultaneously rotate wrist and close hand) [18]. Classifiers can be limited by allowing the user only a predetermined, fixed number of movement types.

Continuous decoders provide another approach to intuitively control a prosthesis. Although nomenclature varies in the literature, here we refer to a continuous decoder as a decoder that predicts kinematic positions anywhere within some movement window (i.e., an infinite number of positions) and learns from continuous kinematic values, not classes with predefined kinematic values. We choose not to refer to continuous decoders as regression-based because not all continuous decoders use regression [24], [25]. Instead of distinct classes of movements, continuous decoders allow independent and simultaneous movement of individual degrees-of-freedom (e.g., finger or wrist movements), allowing the user to potentially produce any grip pattern. Continuous decoders have been used to provide prosthesis control with linear [26]–[28] and nonlinear [29]–[31] algorithms. Nonlinear algorithms generally take longer to train and are subject to overfitting; however, they may capture the nonlinear nature of multi-dimensional EMG data [32], especially for complex movement patterns. Recently, hybrid decoders have combined classification and regression control with promising results [33].

Kalman filters, and related variations, provide a linear approach to continuous control that has been used with cortical [34] and peripheral [26], [35] recordings. Multi-layer perceptrons (MLPs), a relatively simple artificial neural network, have been used for both classification [32], [36], [37] and continuous [29], [38]–[40] control. Convolutional neural networks (CNNs), which, in the prosthetic domain, convolve temporally over their input, have also been used for classification [41], [42] and continuous [30], [31] control. CNNs temporal convolution enables them to learn features from raw EMG data [31], [41], whereas other strategies typically use time-domain features, such as the mean absolute value of segmented EMG.

Beyond the algorithm itself, the data used to train the algorithm affects prosthesis control. Training an algorithm in multiple arm positions can improve prosthesis control whether the position data is used to train the algorithm [32], [43] or not [44], although incorporating positional data may not benefit linear algorithms [45]. For continuous controllers without a discrete set of classes, using training data with simultaneous movements of more than one degree-of-freedom may expose nonlinearities in EMG data and preferentially benefit nonlinear algorithms with higher learning capacities.

Modifying a continuous decoder’s output can improve prosthesis control by improving stability. Smoothing filters, such as a traditional low-pass filter can smooth the output and reduce unwanted jitter [46]. One study incorporated a camera system to selectively low-pass filter the output when a grasp was detected [47]. Low-pass filters inevitably introduce delay into the control. Nonlinear smoothing filters can reduce small-amplitude jitter without slowing larger movements [48], [49].

Prosthesis control improvements are generally demonstrated sequentially in three domains: offline, online, and real-world. Offline comparisons measure the ability of a decoder to map EMG inputs to kinematic outputs, without the user in the loop. Several offline comparisons have helped improve the state of the art for both continuous [40] and classification [17], [18], [32], [42], [50]–[52] control. Although they are a useful first step in improving control, offline improvements do not necessarily imply online improvements [53], largely because they do not incorporate the continuous interaction of the user with the controller. Online improvements incorporate user feedback and have been demonstrated through Fitts’ law or virtual target tasks. Several online comparisons have demonstrated differences among control strategies [28], [30], [31], [36], [41], [53]–[57]. The relationship of online performance to real-world performance is debated [58]–[60]. Comparing decoders through real-world activities of daily living provides the most realistic approximation of how a decoder would function in everyday life. Real-world comparisons have demonstrated the benefits of continuous [61], [62] and classifier [20], [49], [63]–[66] control strategies over commercial control; however, only a couple comparisons have been made among research-grade control strategies [49], [66], both of which were classifiers.

In this work, we conduct a two-phase real-world comparison of 12 continuous decoders that span the three major components affecting control (training data, algorithms, and smoothing). In the first phase, we compare training an algorithm with individual finger or wrist movements vs. combination movements (e.g., simultaneous flexion of all fingers to make a functional grasp) and compare the effect of using a nonlinear smoothing filter [48]. We study these conditions in a two-degree-of-freedom decode using advanced, previously published, continuous control decoders: a modified Kalman filter (mKF) [35], an MLP [29], and a CNN [30],)—all variations together yield 12 different decoders for comparison. In the second phase, we performed a more extensive evaluation of five-degree-of-freedom decodes with the mKF, MLP, and CNN using the combination training and smoothing filter found most beneficial in phase one. We focus on both performance-based objective and patient-centric subjective measures. To our knowledge, this study provides the first comparison of the same individuals using multiple continuous decoders and a physical prosthesis to complete real-world tasks. A large part of our motivation for this study is to identify the “best” decoder among the advanced decoders we have previously published, in part because the results of this study will guide our clinical translation of advanced prosthesis control in a long-term take-home clinical trial.

## Methods

### Signal Acquisition & Prosthesis Setup

Signal acquisition has been described previously [35]. In brief, surface EMG (sEMG) was collected from a sEMG sleeve [67] with the 512-channel Grapevine System (Ripple Neuro LLC, Salt Lake City, UT). Thirty-two single-ended channels were acquired at 1 kHz and filtered with a 6^th^-order high pass Butterworth filter (15 Hz), 2^nd^-order low-pass Butterworth filter (375 Hz), and 60, 120, and 180 Hz notch filters. After connecting the sEMG sleeve to the acquisition device, channels were manually inspected and removed if shorted channels were detected (generally less than two channels). The differential pairs of all monopolar channels were calculated, and features (single-ended and differential) were created at 30 Hz using the mean absolute value of a 300-ms buffer (i.e., 528 features from an overlapping 300-ms boxcar filter). At 30 Hz, the buffer is updated every 33 ms, resulting in a 266-ms overlap between features. This update rate and buffer length has been used by our group extensively with various decoders [30], [35].

Non-amputee participants donned the DEKA LUKE Arm [1] with a bypass socket enabling non-amputee use of a prosthetic arm, which has been shown to be representative of use by individuals with amputation [68].

The prosthesis, bypass socket, and sEMG sleeve are shown in Fig. 1a. The bypass socket, which has a freely-rotating wrist attachment, had rubber bands providing wrist rotation resistance (0.34 Nm to 0.85 Nm at rest and maximal rotation, respectively). Baseline EMG activity was subtracted from each feature before collecting training data by subtracting the average EMG activity during a 10-s period where the user supported the weight of the bypass socket and DEKA LUKE Arm in a neutral position approximately waist height. Training data were recorded as participants mimicked the pre-programmed movements of the DEKA LUKE Arm. The kinematics from the pre-programmed movements were used to fit the algorithms described in the next section.

**Fig. 1.**
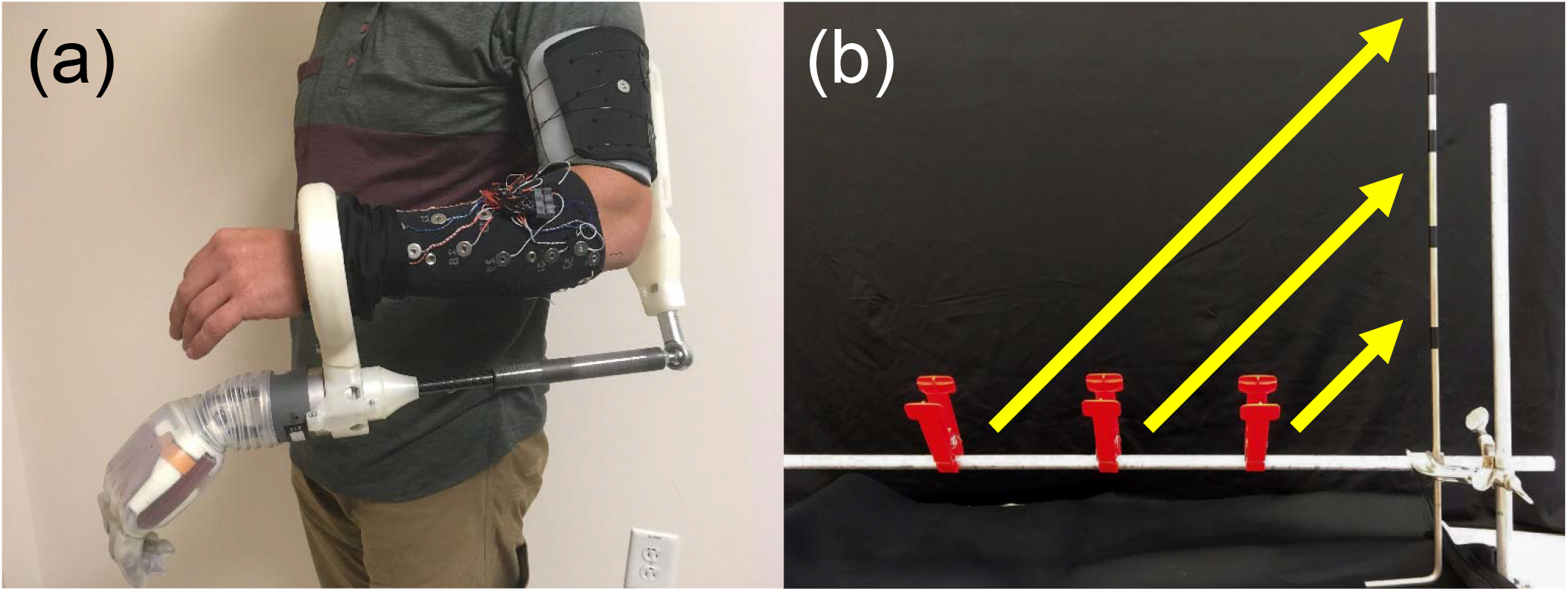
Prothesis setup and phase one task **(a)** The bypass socket and sEMG sleeve enable non-amputees to complete real-world tasks with prosthetic systems. **(b)** The clothespin relocation task. The clothespin relocation task tests prosthesis control with simultaneously active grasping and wrist movements, typical of many activities of daily living.

### Decode Algorithms

The mKF was implemented as has been demonstrated previously [35]. A Gram-Schmidt orthogonalization algorithm [69] was used to select 48 features as inputs for the mKF. Using 48 features was empirically found to be an effective number in [48]. Prior to selecting features, we temporally aligned the kinematics and features by maximizing the average correlation between every feature and kinematic. The 48 EMG features and kinematics were used to fit the parameters of a linear Kalman filter [25]. The **post-hoc** thresholds which were used to modify the Kalman filter online [35], were optimized offline [35]. The feature selection, Kalman filter fitting, and threshold optimization were completed using all the training data (i.e., no training and validation split). The mKF was trained, and the threshold optimized using MATLAB 2018b on a Windows 10 computer with 64 GB RAM and an Intel Xeon E5-2620v3. The same computer was used for runtime predictions.

The nonlinear MLP was implemented as has been demonstrated previously [29]. The MLP consisted of two hidden layers, each with 128 nodes, and a hyperbolic tangent (tanh) activation layer. The training and testing data were split 80% and 20%, respectively. As with the mKF, the MLP used 48 input features as chosen by the same stepwise Gram-Schmidt orthogonalization algorithm to provide cleaner feature data. Prior to selecting features, we temporally aligned the kinematics and features by maximizing the average correlation between every feature and kinematic. The MLP trained for three dataset aggregation iterations, each with ten epochs (found sufficient in [29]). The MLP was trained in Python 3.6 using TensorFlow 1.13 on a Windows 10 computer with 64 GB RAM and an Intel Xeon E5 1650v3 (not trained on a GPU). The same computer was used for runtime predictions.

The nonlinear CNN was implemented as has been demonstrated previously [30]. The CNN, comprising eight total layers, used all 528 features to fit the kinematic values. 75% of the data was used to train the algorithm, and the remaining 25% was used as a validation set. The network trained until the loss on the validation set was equal to or larger than the previously smallest loss for five iterations. The CNN was trained using MATLAB 2018b on an NVIDIA Quadro M4000 8 GB graphics card. The system ran on Windows 10 and had 128 GB RAM and an Intel Xeon E5-2620v4 processor. The trained network was transferred to a Windows 10 computer with 64 GB RAM and an Intel Xeon E5-2620v3 processor for runtime predictions.

After training, users were given position control of the prosthesis. In position control, the prosthesis returns to its rest position if the user is not actively making a movement. In velocity control (the counterpart of position control), the prosthesis does not move when the user rests. For tasks requiring multiple active movements, position control forces the user to complete both movements simultaneously, whereas velocity control allows the user to complete these movements sequentially.

### Smoothing Filter

A smoothing filter was implemented as has been demonstrated previously [48]. The “latching filter” we used is a computationally inexpensive, recursive, nonlinear filter that smooths small-amplitude jitter but allows quick changes to its output. The latching filter nonlinearly adjusts its level of smoothing based on the difference between the previous, smoothed decoder estimate and the current decoder estimate to determine how much to move in the direction of the current decoder estimate (i.e., how much to smooth the transition from the previous estimate to the new estimate). For small estimate changes, the output is heavily smoothed, resulting in stability for low-amplitude movements. For large estimate changes, the output is nearly unsmoothed, resulting in quick movements. The latching filter makes the reasonable assumption that jitter amplitude is small relative to the movement range and that jitter occurs more frequently than do intended small movements.

### Study Overview

We conducted this study in two phases (Fig. 2a). In the first phase, we compared training paradigms and the use of a smoothing filter for all three algorithms, totaling 12 decoders: three algorithms, two training paradigms, and two output types. Due to the number of decoders and the consequential experiment duration, we selected a simple task, the clothespin relocation task, with a low, two degree-of-freedom control strategy (grasp and wrist rotation), which simplified the training and allowed the experiments to be completed in 2-3 hours.

**Fig. 2.**
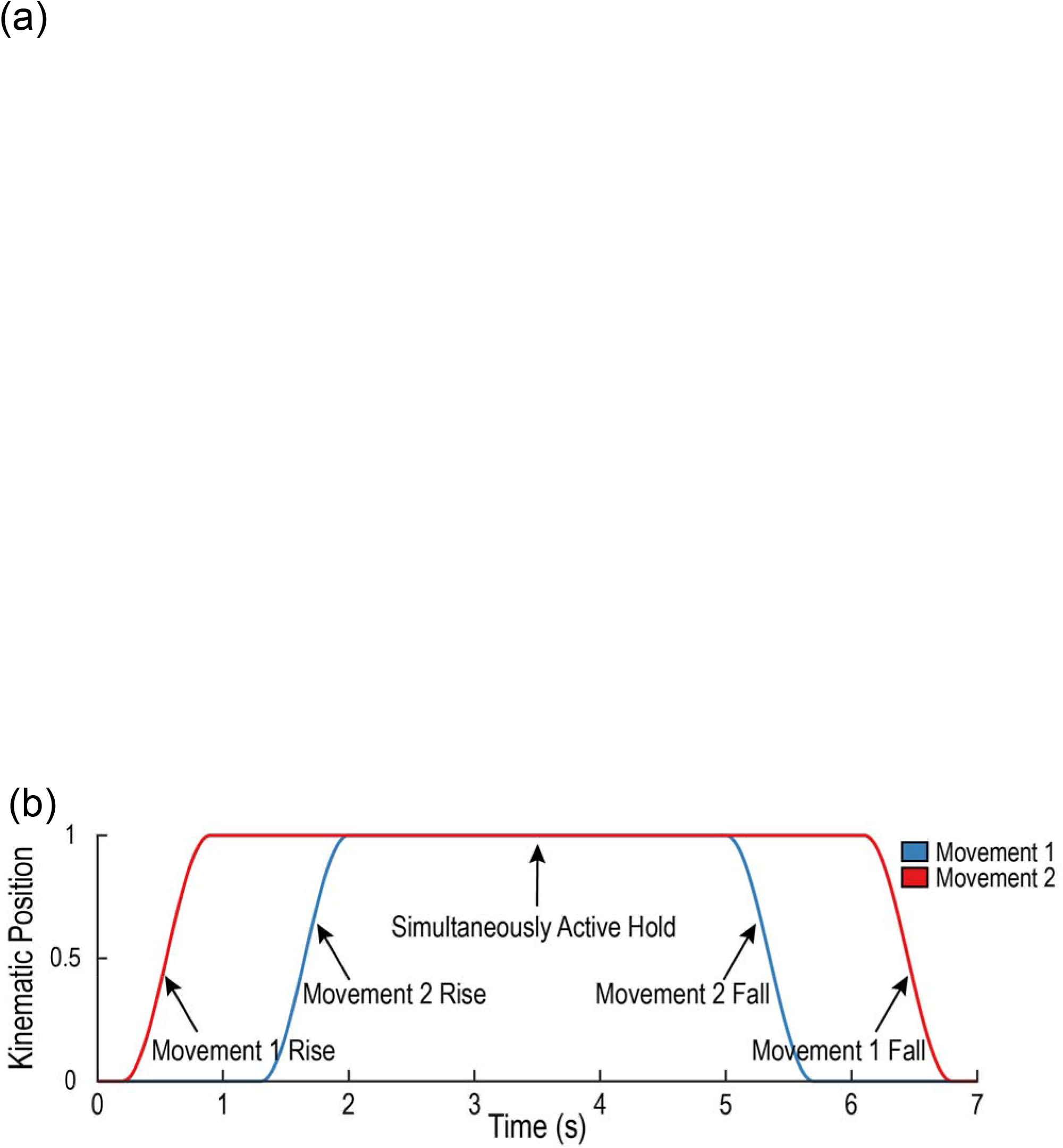
**(a)** Overview of the two-phase functional comparison of 12 prosthesis control strategies. In phase one, we compared collecting training data with individual movements vs. combination movements and compared using raw algorithm output vs. nonlinear output smoothing with a latching filter for an mKF, MLP, and CNN in a clothespin relocation task. In phase two, we compared the mKF, MLP, and CNN with fragile egg, zipping, pouring, and folding tasks. **(b)** Example combination movement, where two degrees-of-freedom are simultaneously active during the middle portion of the trial.

In phase two, we increased the decoder output to five degrees-of-freedom to evaluate the three algorithms’ control in a high-degree-of-freedom setting representative of modern myoelectric prostheses. The degrees-of-freedom used were: (1) thumb flexion, (2) index finger flexion, (3) middle, ring, and little finger flexion (which are mechanically coupled on the DEKA LUKE Arm), (4) wrist flexion, and (5) wrist rotation. We used the results of the first phase to inform the training paradigm and smoothing filter use. As we were comparing only three algorithms in the second phase, we were able to perform four tasks (fragile egg, zipper, pouring, and folding) in a 2-3-hour experimental session.

### Phase One: Training Type and Output Smoothing Comparison

#### Training Paradigms

Ten participants (24 ± 4 years [mean ± SD]; seven right-handed, three left-handed; nine male, one female) with no prior prosthesis experience mimicked the DEKA LUKE Arm movements in three training periods to provide training data for the control algorithms. The two-degree-of-freedom decode training consisted of opening and closing the hand as well as rotation of the wrist. Prior to each training period, study administrators coached the participants through one trial of each movement, to acquaint the participants with mimicking the movements. In the first training period, users completed 12 trials of individual movements: close hand, open hand, pronate, supinate. Each individual movement consisted of a 0.7-s rise time, 3-s hold, and 0.7-s return to rest position (for a total of 4.4 s per movement). The second training period repeated the first period; however, only four trials of each movement were completed. The first and second training periods took 5 and 1.5 minutes, respectively.

The third period introduced combination movements, where both degrees-of-freedom were simultaneously active for the middle portion of the trial (see Fig. 2b for a visual representation). Users completed four trials each of eight different movements: close hand and pronate, close hand and supinate, open hand and pronate, open hand and supinate, pronate and close hand, supinate and close hand, pronate and open hand, and supinate and open hand. For one combination movement, the first degree-of-freedom moved to its intended position with a 0.7-s rise time, 5.2-s hold, and 0.7-s return to resting position. The second degree-of-freedom moved to its intended position 0.4 s after the first degree-of-freedom reached its intended position with a 0.7-s rise time, 3.0-s hold, and 0.7-s return to its rest position. In total, one combination training trial consisted of 6.6 s. The third training period took 5 minutes.

To compare individual and combination training paradigms, the training sets were merged into two sets of equal duration before being used to train the decoder (so that any improved performance would not be due to the decoder training on more data). The first and second training periods were combined to produce an individual-movement-only set. The second and third training periods were combined to produce a set with individual and combination movements. After merging the datasets, the mKF, CNN, and MLP were trained on the kinematic and EMG data for proportional control.

#### Modified Clothespin Relocation Task

In the modified clothespin relocation task (Fig. 1b), participants transferred three large clothespins from a horizontal bar to a vertical bar on the side of the dominant hand, referred to as the “inside-out” task [70]. The task was chosen for several reasons: its simplicity allows the user to quickly learn and master the task, mitigating effects due to learning; its short duration allows several trials to be completed rapidly, which was necessary for comparing 12 decoders; and, it necessarily requires simultaneously active degrees-of-freedom, typical of many daily activities. We have used this task with the DEKA LUKE Arm previously [71].

Participants started a timer with the prosthetic hand, transferred a single clothespin, and stopped the timer with the prosthetic hand. A trial was considered failed when a clothespin was dropped. Participants were instructed that the success rate (proportion of successfully transferred clothespins) and the transfer time would be recorded. There was no time limit for the task. Participants completed nine trials (three sets of three) with each decoder. The order of the control paradigm was randomized among subjects to prevent an order effect. After completing nine trials with a particular decoder, participants completed the NASA Task Load Index (TLX) subjective workload survey [72].

Participants completed the task for 12 total decoders, including each possible combination of training paradigm (individual and combination), algorithm (mKF, MLP, and CNN), and output type (raw and smoothed).

#### Analysis

We compared the effect of training paradigm and smoothing filter for each algorithm with α = 0.05. The data were significantly non-normal for several conditions (Shapiro-Wilk test [73]), so nonparametric statistics were used. Median completion times for successful trials were calculated for each condition for each participant (N = 10 participants). The number of drops was calculated for each condition for each participant. For the training paradigm comparison, the data were aggregated to compare individual and combination trainings with Wilcoxon’s signed-rank test [74] for each algorithm; each group contained both smoothed and raw output types (N = 20; two medians for each participant). For the output type comparison, the data were aggregated to compare raw and smoothed outputs with Wilcoxon’s signed-rank test for each algorithm; each group contained both individual and combination trainings (N = 20; two medians for each participant). Because the hypotheses were pre-planned, no correction for multiple comparisons was applied [75].

We compared the extent that the algorithms predicted simultaneously active degrees-of-freedom. Because this was a *post-hoc* analysis, we did not have digital markers indicating when a clothespin trial was started. Instead, we examined the proportion of time an algorithm predicted hand movement and wrist rotation simultaneously, divided by the total time the hand or wrist was active. We determined the threshold for “active” movements by the range of movements needed to complete the clothespin task. The wrist needed to rotate a minimum of 35% of its movement window to place the clothespin, and the hand needed to close a minimum of 20% of its movement window to hold the clothespin. We counted only simultaneous movements over 1 s in duration to exclude potentially sporadic predictions that were likely not part of the task completion. We used the median proportion of time each algorithm predicted simultaneous movements for the algorithm comparisons. The medians across participants were distributed normally (Shapiro-Wilk test [73], so we conducted a one-way ANOVA [76] to determine if there were significant differences among the algorithms. If the ANOVA revealed differences, individual comparisons were made using Tukey’s Honestly Significant Difference procedure [77].

### Phase Two: Algorithm Comparison

#### Training Paradigm

In the second phase, we extended the degrees-of-freedom from two to five to assess the algorithms’ performance in more complex, biologically realistic conditions. Users controlled the following degrees-of-freedom: flexion/extension of the thumb, flexion/extension of the index finger, flexion/extension of the middle, ring, and little fingers (which are mechanically coupled on the DEKA LUKE Arm), flexion/extension of the wrist, and pronation/supination of the wrist. Study administrators coached the participants through a single trial of each movement type before each training period. The training for phase two consisted of four distinct training periods, each 5 minutes long, for a total of 20 minutes.

Nine participants (30 ± 14 years [mean ± SD]; seven right-handed, two left-handed; seven male, two female) with some prior prosthesis experience (2-3 hours) mimicked the prosthesis through movements of individual degrees-of-freedom. Six participants from phase one had completed the phase one study; however, not all the previous participants were able return for the phase two study. The remaining three had completed other prosthesis studies with our group. Movements consisted of a 0.7-s rise time, 5.2-s hold time, 0.7-s fall time (for a total of 6.6 s per movement). Participants mimicked four trials on each degree-of-freedom. For the second through fourth training periods, combination movements were introduced.

The second period consisted of grasp movements (simultaneous flexion of the thumb, index, and middle, ring, and little degrees-of-freedom) combined with wrist movements (flexion, extension, pronation, and supination). Participants completed the first grasp movement alone, with a 0.7-s rise time, 5.2-s hold time, and 0.7-s fall time (6.6 s total). Participants then completed grasp and wrist combination movements, where both the grasp and wrist movement were simultaneously active in the middle portion of a trial (Fig. 2b). In these combination trials, the grasp movement rose to its intended position in 0.7 s, held for 5.2 s, and returned to its original position in 0.7 s. The wrist movement started 0.4 s after the first movement reached its intended position, moved to its intended position with a 0.7-s rise time, held for 3.0 s, and returned to its original position in 0.7 s. These combination movements were completed for each wrist movement (wrist flexion, extension, pronation, and supination). Then, they were completed in the reverse order, where the wrist movement occurred first, and the grasp movement occurred part-way through the wrist movement. We chose to complete the movements in both orders due to the CNN convolving over time. Participants completed four trials of each sequence, resulting in 36 total trials.

Periods three and four repeated the second period; however, the grasping movement was replaced with opening the hand (extending the thumb, index, and middle, ring, and little digits) and a pinch movement (flexing thumb and index digits), respectively. After merging the datasets from the four training periods, the mKF, CNN, and MLP were trained on the kinematic and EMG data for proportional control.

#### Activities-of-Daily-Living

In the second phase, participants completed four tasks representative of activities of daily living in the following order: moving a fragile egg, using a zipper, pouring, and folding a towel (Fig. 3). A time limit was specified for each task. The order of the three decode algorithms (mKF, CNN, and MLP) was randomized for each participant to avoid order effects, with the same random order being used within each of the four tasks. After completing a task with each algorithm, participants ranked the algorithms in order of preference. Examples of the tasks are included in the supplementary material (Additional file 1: Video 1). Each task is now discussed in greater detail.

**Fig. 3.**
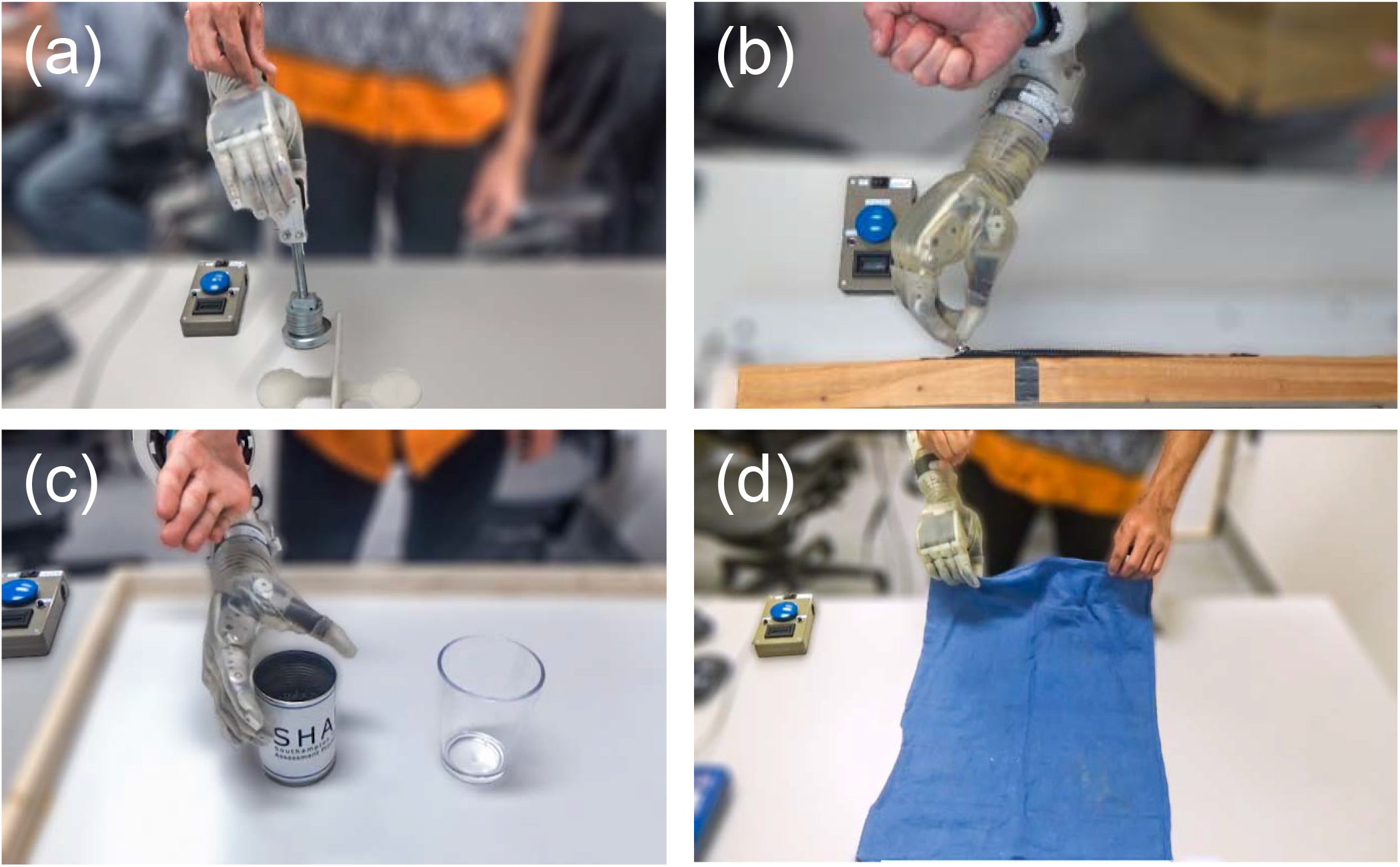
Phase two: Activities of daily living represent realistic prosthesis use. **(a)** The fragile egg task tests precision control and tests the proportionality of a decode algorithm. **(b)** The zipper task requires stable pinching and simultaneous wrist rotation. **(c)** The pouring task requires stable grasping with smooth wrist rotation. **(d)** The folding task requires two-handed object manipulation and stable grasps during positional changes.

#### Fragile Egg Task

In the fragile egg task, participants moved a mechanical “fragile egg” [78], [79] horizontally 15 cm from one side of a 6.25-cm vertical barrier to the other within a 45-s time limit, beginning on the side of the dominant hand. This task evaluated the dexterity provided by each decode algorithm in that if too much force was applied, the mechanical egg would “break” and emit an audible click. The participants were instructed to use a two-finger pinch to pick up the mechanical egg. The breaking force for these experiments was set at about 20 N, and the device mass was about 615 g (0.03 N/g).

For each trial, participants started a timer with the prosthetic hand, transferred the mechanical egg, and stopped the timer with the prosthetic hand. A trial was considered failed if the mechanical egg was broken or dropped. Participants were instructed that both the success rate (the proportion of trials in which transfer was successful) and the transfer time would be recorded. When introduced to the task, participants were given two minutes of practice with each decode algorithm to decrease the learning effects in the recorded trials. Participants attempted 15 trials with each decode algorithm. Immediately before the recorded trials commenced, participants were given one final practice trial with the given decode algorithm. A verbal warning alerted the participants when there were 15 s remaining. After attempting the 15 recorded trials with a particular algorithm, participants completed a NASA-TLX survey.

#### Zipper Task

In the zipper task, participants closed a horizontally mounted zipper within a 30-s time limit, beginning on the side of the dominant hand. Timed-out trials were reported as 30 s. The zipper task tests precision grasping with combination wrist movements. For a trial to be successful, the zipper must have closed by at least 6.5 cm, as indicated with a mark. We modified the base of the zipper so that the handle would not lie less than 45 degrees from being flat because the shape of the prosthetic fingers prevented gripping the zipper when lying flat. For each trial, participants started a timer with the prosthetic hand, closed the zipper, and stopped the timer with the prosthetic hand. Participants were instructed that both the number of releases (times the zipper slipped from their grasp) as well as the time would be recorded. When introduced to the task, participants were given three practice trials with each decode algorithm in order to decrease the learning effects in the recorded trials. Participants attempted five trials with each decode algorithm. Immediately before the recorded trials commenced, participants were given one final practice trial with the given decode algorithm. After attempting the five recorded trials with a particular algorithm, participants completed a NASA-TLX survey.

#### Pouring Task

In the pouring task, participants poured rice from an aluminum can into a cup within a 1-min time limit. The rice-filled can was placed on the side of the dominant hand, and the empty cup was placed near the midline. This task assessed the ability of the decode algorithms to maintain a grasp during wrist rotation. For these experiments, the aluminum can was filled with 200 g of rice.

For each trial, participants started a timer with the prosthetic hand, poured the rice from the can (7.6 cm diameter, 11.3 cm tall) to the cup (8.6 cm diameter, 11.7 cm tall), and stopped the timer with the prosthetic hand. Participants were instructed that the amount of rice (in grams) successfully transferred to the cup as well as the transfer time would be recorded. The participants were not allowed to use their native hand to stabilize the cup, and if the can was dropped the trial was considered unsuccessful. When introduced to the task, participants were given three practice trials with each decode algorithm in order to decrease the learning effects in the recorded trials. Participants attempted five trials with each decode algorithm. Immediately before the recorded trials commenced, participants were given one final practice trial with the given decode algorithm. After attempting the five recorded trials with a particular algorithm, participants completed a NASA-TLX survey.

#### Folding Task

In the folding task, participants folded a small hand towel (39 cm by 58 cm) twice using a pinch grasp within a 1-min time limit. The first fold was done from short-end to short-end. The participant could then use their non-dominant native hand to rotate the towel 90 degrees for the second fold, which was also from short-end to short-end. For both folds, participants were instructed not to use the edge of the table to facilitate grasping the towel. Additionally, they were not to use their native hand to guide the end of the towel into the prosthesis. Ideally, participants simultaneously grasped the short edge of the towel with their native hand and the prosthesis.

For each trial, participants started a timer with the prosthetic hand, folded the towel twice, and stopped the timer with the prosthetic hand. Participants were instructed that the number of releases (times the towel slipped from their grasp) and the time would be recorded. When introduced to the task, participants were given three practice trials with each decode algorithm to decrease the learning effects in the recorded trials. Participants attempted five trials with each decode algorithm. Immediately before the recorded trials commenced, participants were given one final practice trial with the given decode algorithm. After attempting the five recorded trials with a particular algorithm, participants completed a NASA-TLX survey.

#### Analysis

We compared algorithms for each metric within each task with α = 0.05. The data were significantly non-normal for several conditions (Shapiro-Wilk test [73]), so nonparametric statistics were used. Median completion times for successful trials were calculated for each condition for each participant (N = 9 participants). For the pouring task, the task was considered successful if the total amount transferred was greater than 199.9 g (no spill). For each metric, a Kruskal-Wallis test [80] was used to determine if there were differences among the decode algorithms. If the Kruskal-Wallis revealed differences, paired comparisons were made using Wilcoxon’s signed-rank test [74]. Because the hypotheses were pre-planned, no correction for multiple comparisons was applied [75]. *Post-hoc*, we decided to compare aggregate user preferences. We grouped the user preferences for each task and performed a Kruskal-Wallis test across algorithms. As this was an unplanned comparison, we corrected the pairwise comparisons with the Dunn-Sidak correction for multiple comparisons [81].

## Results

### Phase one: Comparing Latching Filter and Training Type

#### Latching Filter Comparison

The latching filter generally reduced the number of dropped clothespins but sometimes increased the clothespin transfer time (Fig. 4a-c). For the MLP, dropped clothespins were significantly reduced from 3 to 1 (p < 0.01; Fig. 4a). For the CNN, dropped clothespins were significantly reduced; however, the median values were both 1 (p < 0.05; Fig. 4a). The latching filter significantly decreased transfer times from 11.0 s to 7.4 s for the MLP (p < 0.01), but significantly increased transfer times for the CNN and mKF from 6.9 s to 7.2 s (p < 0.05) and 7.8 s to 10.1 s (p < 0.001), respectively (Fig. 4b). The latching filter significantly reduced subjective workload for the MLP from 54 to 41 (p < 0.05) but had no significant effect on subjective workload for the CNN and mKF (Fig. 4c).

**Fig. 4.**
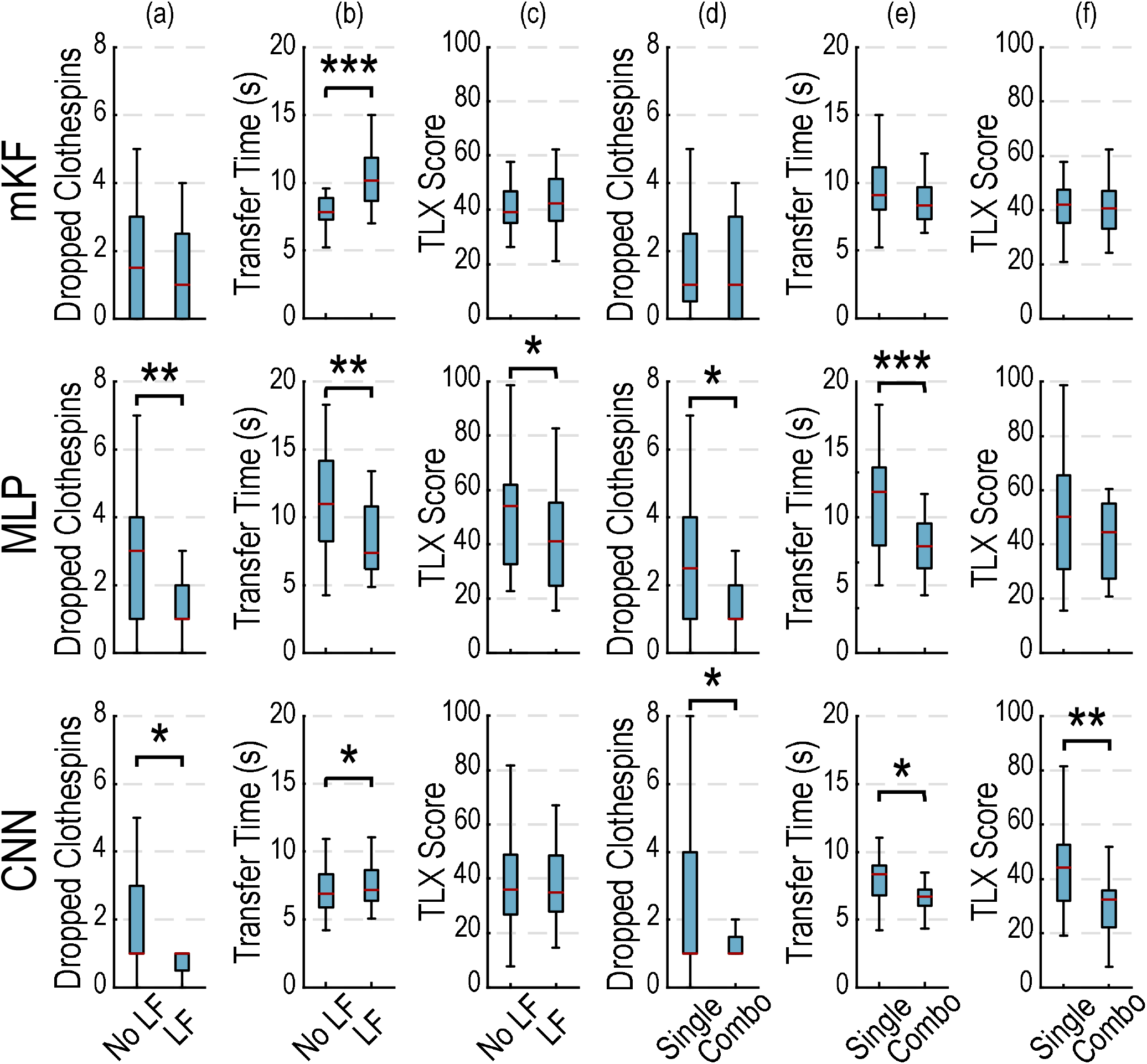
Phase one: The latching filter (LF) improves prosthesis control, sometimes at the cost of speed, and combination training improves control accuracy and speed. **(a)** The latching filter significantly reduced dropped clothespins for the MLP and CNN. **(b)** The latching filter significantly increased transfer time for the mKF and CNN, and significantly reduced transfer time for the MLP. **(c)** The latching filter significantly reduced the subjective workload for the MLP, evidenced by a lower NASA Task Load Index (TLX) score. **(d)** Combination training significantly reduced dropped clothespins for the MLP and CNN. **(e)** Combination training significantly reduced transfer times for the MLP and CNN. **(f)** Combination training significantly reduced the subjective workload for the CNN, evidenced by a lower NASA Task Load Index (TLX) score. In these comparisons, the variable being compared contains aggregate data from the variable not being compared (e.g., in LF comparisons, each group contains both single and combination training types). Boxplots show median (red line), inter-quartile range (blue box), and most extreme, non-outlier values (1.5 x inter-quartile range; black whiskers).

#### Training Type Comparison

Training with combination movements generally improved the outcomes for the clothespin relocation task (Fig. 4d-f). Training with combination movements significantly reduced dropped clothespins for the MLP from 2.5 to 1 (p < 0.05; Fig. 4d). For the CNN, dropped clothespins were significantly reduced; however, the median values were both 1 (p < 0.05; Fig. 4d). Training with combinations significantly reduced the median time needed to transfer a clothespin for CNN and MLP from 8.3 s to 6.7 s (p < 0.05) and 11.8 s to 7.9 s (p < 0.001), respectively (Fig. 4e). Combination training significantly reduced the subjective workload for the CNN, from 44 to 32 (p < 0.01; Fig. 4f). Training type had no significant effect on the subjective workload for the mKF and MLP; however, both were reduced with combination training (Fig. 4f).

#### Simultaneous Movement Analysis

Simultaneous movements were significantly above zero for all three decoders (p < 0.001, all). The proportions of simultaneously active degrees-of-freedom were significantly different between algorithms (p < 0.01). The MLP had significantly more simultaneous movement than the CNN (16.6% ± .5% and 9.0% ± 0.6%, respectively [mean ± SEM]; p < 0.01). The mKF had significantly more simultaneous movement than the CNN (15.6% ± 0.5% and 9.0% ± 0.6%, respectively [mean ± SEM]; p < 0.05).

### Phase Two: Comparing Algorithms

#### Fragile Egg Task

Preferences for the fragile egg task were significantly different for the different decode algorithms (p < 0.01; Fig. 5e). The CNN was significantly preferred over the MLP (median ranks 1 and 3, respectively; p < 0.05). Similarly, the mKF strongly trended toward being preferred over the MLP (median ranks 2 and 3, respectively; p = 0.05). Subjective workload also significantly differed among algorithms (p < 0.05; Fig. 5b). The CNN had significantly lower workload than did the MLP (51, 74, respectively; p < 0.01). The mKF had significantly lower workload than did the MLP (51, 74, respectively; p < 0.05). There were no significant differences in success rate across the algorithms; however, the mKF and CNN were generally more successful than the MLP (Fig. 5c). There were no significant differences in completion time across algorithms, although the mKF generally performed faster than the CNN and MLP (Fig. 5a).

**Fig. 5.**
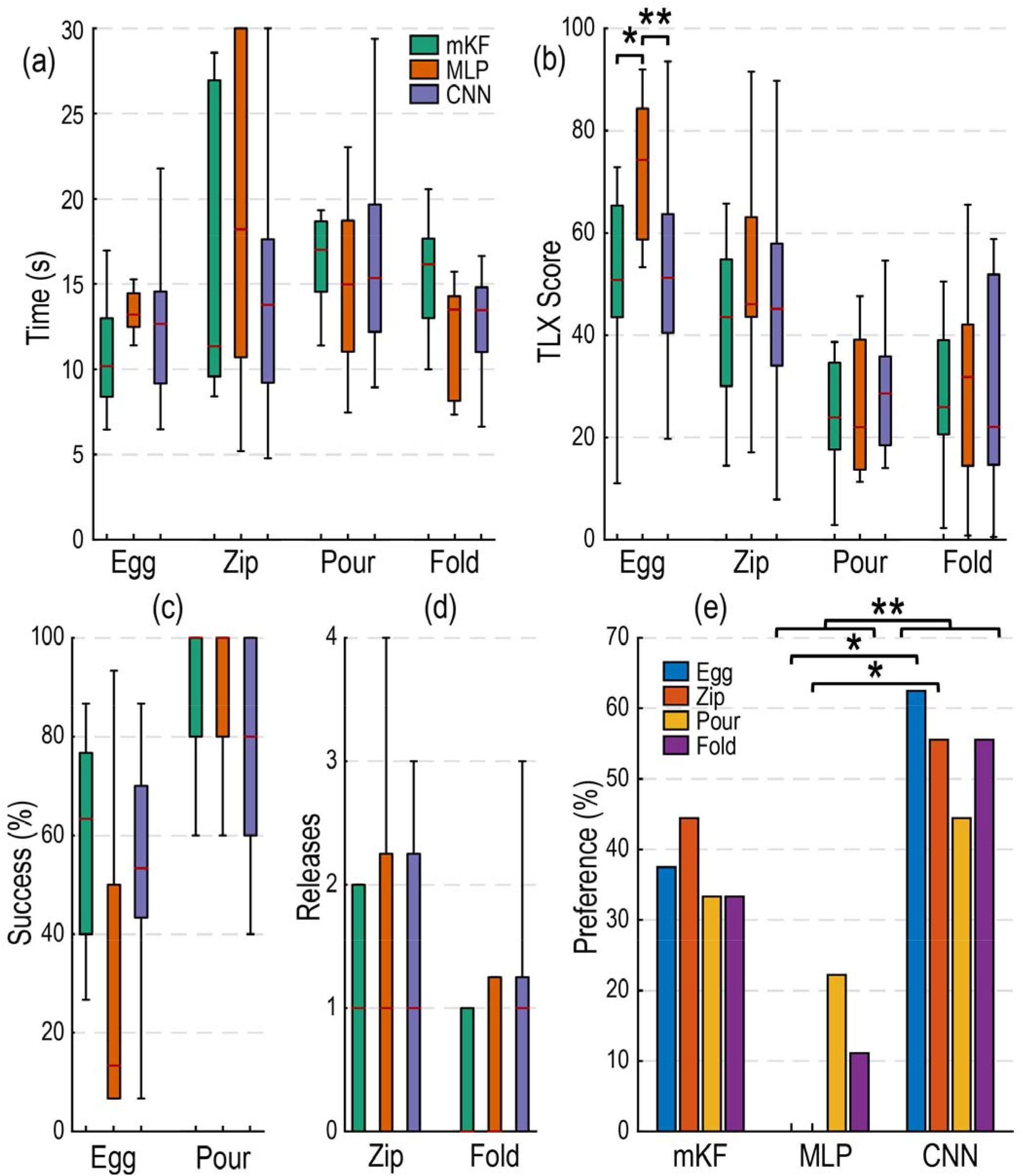
Phase two: subjective user-focused metrics reveal differences that objective performance-based measures may not. **(a)** Completion times did not significantly differ for any of the tasks. The mKF was generally faster for tasks perceived as more difficult, and slower for tasks perceived as less difficult. **(b)** The mKF and CNN were significantly easier to use than the MLP for the fragile egg task, as evidenced by a lower NASA Task Load Index (TLX) score. **(c)** mKF and CNN outperform MLP in fragile egg task, although the difference was insignificant. All algorithms had high success in the pouring task. **(d)** All algorithms performed well and did not significantly differ in the zipping and folding tasks. **(e)** The CNN was significantly preferred over the MLP, aggregated across tasks. The mKF trended towards significance over the MLP across all tasks. The CNN was significantly preferred over the MLP for the fragile egg and zipper tasks. Boxplots show median (red line), inter-quartile range (colored boxes), and most extreme, non-outlier values (1.5 x inter-quartile range; black whiskers).

#### Zipper Task

Preferences for the zipper task differed significantly among algorithms (p < 0.05; Fig. 5e). The CNN was significantly preferred over the MLP (median ranks 1 and 3, respectively; p < 0.05). There were no significant differences in completion time (Fig. 5a), but the median completion time for the mKF was the fastest. Inadvertent releases and subjective workload for the decodes did not significantly differ among algorithms (Fig. 5d, 5b, respectively).

#### Pouring Task

The pouring task was generally perceived as less difficult than the fragile egg and zipper tasks (Fig. 5b). There were no statistical differences for any of the recorded metrics (time, success rate, preference, and subjective workload). The MLP was preferred by a few participants, in contrast to the fragile egg and zipper tasks where it was never preferred (Fig. 5e). Participants had high success rates for the pouring task across algorithms and rarely spilled (Fig. 5c); we report only on whether the task was completed without spills or drops. Completion times were similar among algorithms (Fig. 5a).

#### Folding Task

The folding task was generally perceived as less difficult than the fragile egg and zipper tasks (Fig. 5b) and did not result in any significant differences for the recorded metrics. More participants seem to prefer the CNN for this task (Fig. 5e) even though the trend suggests they inadvertently released the towel more than with the mKF or MLP. The CNN and MLP may also perform faster than the mKF (Fig. 5a).

#### Aggregate Preferences Across Tasks

Aggregated across tasks, user preferences were significantly different (p < 0.05; Fig. 5e). Users preferred the CNN over the MLP (median ranks 1 and 3, respectively; p < 0.01), and the mKF trended toward being preferred over the MLP when aggregated across tasks (median ranks 2 and 3, respectively; p = 0.07).

#### Algorithm Training and Prediction Times

On average, the mKF trained in 0.7 s, the MLP trained in 47 s, and the CNN trained in 135 s in phase one. The threshold optimization averaged 155 s in phase one. In phase two, the mKF trained in 2.7 s, the MLP trained in 193 s, and the CNN trained in 262 s. The threshold optimization took 480 s in phase two. For testing, the mKF took 0.6 ms, the MLP took 1.4 ms, and the CNN took 3.0 ms. The latching filter took 0.027 ms to modify the algorithm output. Detailed training and testing results are included in Table 1.

**Table 1.** Overview of Training and Prediction Times

| Decoder Component | Algorithm Training |  |  | Testing |
| --- | --- | --- | --- | --- |
| | Phase One | | Phase Two | Mean ± SD (ms) |
| | Individual<br>Mean ± SD (s) | Combination<br>Mean ± SD (s) | Combination<br>Mean ± SD (s) | |
| mKF | 0.68 ± 0.04 | 0.71 ± 0.05 | 2.68 ± 0.87 | 0.619 ± 0.192 |
| MLP | 45.43 ± 5.15 | 49.32 ± 8.61 | 192.83 ± 6.24 | 1.388 ± 0.151 |
| CNN | 155.97 ± 29.25 | 114.11 ± 37.26 | 261.73 ± 38.00 | 2.953 ± 6.922 |
| Optimized Threshold | 151.33 ± 12.98 | 157.94 ± 19.54 | 480.17 ± 43.68 | N/A |
| Latching Filter | N/A | N/A | N/A | 0.027 ± 0.048 |

## Discussion

In this two-phase study, we demonstrate the importance of using diverse training sets that include combinatorial movements; the value of smoothing the algorithm output; and the efficacy of linear and nonlinear continuous decode algorithms for restoring functional ability to prosthesis users. To our knowledge, no study has attempted to compare real-world functional improvements of continuous prosthesis control using multiple algorithms, training paradigms, and output smoothing from the same participants.

In phase one, we found that training on combination movements improves functional performance and that a latching filter [48] can reduce jitter albeit with some cost in speed. In phase two, we found that a linearly-based decode algorithm (the mKF) can perform similarly to nonlinear deep-learning techniques (the CNN and MLP). From a clinical perspective, the mKF may be preferable sometimes due to reduced algorithm training time and computational resources required for real-time control. Phase two demonstrates how patient-centric outcomes (user preferences and subjective workload) elucidated differences among algorithms when performance metrics alone could not.

Adding combination movements to the training data benefited the participants two-fold: they completed tasks faster and with greater control. The improved speed and control seem to preferentially benefit nonlinear algorithms (MLP, CNN) by producing more consistent control among participants, whereas the mKF was relatively consistent in both conditions. In terms of speed, the MLP benefited the most from the combination movement training; however, it was also the slowest in the single-movement training paradigm and had the most room for improvement. In terms of subjective workload, results from combination training suggest that nonlinear approaches benefited most from combination training; however, the difference was significant only for the CNN. Perhaps combination trainings introduce nonlinearities that preferentially benefit nonlinear neural networks.

The latching filter generally enhanced control at the cost of speed. We were surprised how little the latching filter benefited the mKF relative to the CNN and MLP. The latching filter uses a hyperparameter that adjusts the level of “latching” (and inherent speed) of kinematic changes. The latching filter uses the hyperparameter to preferentially dampen small changes in the decoder estimates (which are assumed to be inadvertent movements) while allowing gross movements to occur with little or no dampening [48]. Our piloting and informal use of these algorithms in lab suggest nonlinear approaches generally predict kinematic positions similar to the data used for training and thus move to heavily trained positions quickly and unimpeded by the latching filter (e.g., rest, full flexion); in contrast, the Kalman filter predicts kinematics better dispersed throughout the kinematic range, which may be due to a relatively low Kalman gain [30] that places more value on past estimates than current measurements and decreases responsiveness. Thus, gross movements could be slowed by a low Kalman gain and the latching filter as they are initiated. We expect that the hyperparameter setting favored the nonlinear algorithms, as raw decoder output from the nonlinear algorithms likely moved between extreme positions (e.g., open hand to closed hand) faster and were not slowed as much by the latching filter when gross movement was intended. We expect that optimizing the hyperparameter could improve speed and control for the mKF especially, but also the CNN and MLP; however, such an optimization is beyond the scope of this study. The lack of improvement in dropped clothespins with the mKF might also have occurred because its control was different than the other two decoders, and so the participants did not become as accustomed to its responsiveness.

In phase two, we examined the decode algorithm’s performance using combination training and the latching filter in a high-degree-of-freedom setting (five; increased from two in phase one). With the reduced experimental condition count in this phase, we were able to thoroughly investigate the algorithms’ performance across a broader range of functional tasks. This phase also included a larger, more diverse training dataset, which we anticipated might preferentially benefit nonlinear algorithms.

We chose tasks representative of daily life that probe qualitatively different aspects of prosthesis control. The fragile egg task, generally judged most difficult by participants, requires precise movements and tests the decoder’s proportionality. The zipper task, also judged difficult by participants, requires consistent precision pinching with simultaneous wrist rotation. The fragile egg tasks and zipper tasks, which test precise movements, could be more difficult if the task were occluded by the bypass socket or prosthesis itself. However, a similar occlusion may occur in the real world. We did not have any participants comment on the difficulty of the tasks as being due to occlusion. The pouring task necessitates smooth wrist rotation during stable grasping. The folding task requires two-handed object manipulation and grasp stability with positional changes which often degrade control performance [59]. We found the variety of the tasks to be useful, but the tasks were easier for the participants than pilot experiments suggested and, in some cases, resulted in floor and ceiling performance effects. The tasks would need to be more difficult to better elucidate performance differences among the algorithms in the future.

In part because of these floor and ceiling effects, user preferences provided strong evidence for identifying the “best” algorithm. Whereas many performance metrics varied only slightly, user preferences showed greater differences among algorithms. Aggregated across tasks, preferences indicated the CNN was preferred over the MLP, and the mKF also trended toward being preferred over the MLP. As testing multiple algorithms in the same experimental sessions allowed users to rank their preferred algorithms, we see incorporating user preferences as a strength of the study. Translating research practices to the clinic requires patient-centric, data-driven approaches. Although performance differences may be small among algorithms, our findings show that differences in user experience exist across the participants, providing additional evidence toward clinical translation. Subjective workload also provided useful information on user experience, showing that the CNN and mKF were easier to use during the fragile egg task. From the user’s perspective, subjective workload may be the difference between adopting or abandoning a prosthesis; while two prosthetic systems may be capable of similar performance, the most intuitive, easy-to-use prosthesis will certainly be preferred.

We found it surprising that the mKF performed similarly to the nonlinear CNN whereas the MLP underperformed. Our informal use of the mKF suggests that the mKF performed well because it has a smoother, more predictable response to input changes, which can improve functional control [53]. We anticipate the CNN performed well because it used all 528 of the EMG features (versus 48 from the channel selection algorithm for mKF and MLP) and convolves across time, thus training on more complex data and potentially learning from temporal characteristics which could be advantageous over the MLP. In [29], the MLP uses the first 16 principal components as input features, different from our use of 48 selected features, which could be another factor in its poorer performance. We informally observed that participants struggled to proportionally control the prosthesis for the MLP, and it would often close quickly and break the fragile egg, which may indicate a lack of generalization from the training data.

It is important to note that the purpose of our study was not to provide an exhaustive comparison of the mKF, CNNs and the MLP per se; rather, our goal was to determine whether combination training sets and the use of a latching filter would improve these previously published decoders, and which combinations to recommend for clinical translation. Although one algorithm may perform better under different circumstances (e.g., the number or type of features provided, training kinematics associated with mirror movements, or the amount of data in testing and validation sets) exploring all possible variations would have required even more subject time, which was already limited. Even with the reported experimental variations, most participants were starting to fatigue and eager to remove the heavy prosthesis by the end of the study—hence there was a practical limit to the number of decoders and variations in our comparison (e.g., only training type and latching filter using previously published algorithms).

The training data used are critical for the performance of any decoder. In our study, we used mimicry training, where the user follows the prosthesis through preprogrammed movements, and the algorithm fits the feature inputs to the kinematics of the prosthesis. Inherent in this training paradigm is the assumption that the user perfectly mimics the kinematics of the hand, which inevitably is an imperfect assumption. Imperfect kinematic and feature data could have been preferentially detrimental to certain decoders (e.g., the MLP may perform better with cleaner labels). Another method for creating kinematics is to use mirror training, where the kinematics of the opposite hand are recorded while the user completes bilaterally mirrored movements. This approach has been shown to provide reliable training data for prosthesis control [82], [83] and could be further studied in real-world environments. Even though we did not use mirror movements, we aligned our kinematics and features during channel selection to improve performance. Additionally, some of the training data used single finger motions not exclusively in use for the tasks. It is possible that the training paradigms could have been improved for the specific tasks completed in this study; however, in real-world settings the tasks are not known *a priori* and therefore a generalizable training dataset is preferable. Recent studies have reported on some of the most used grasp types [84], [85]. Based on the findings of these studies, a specific, yet generalizable, training set could be developed for real-world usage. Another method of improving features could include optimizing electrode placement. Although beyond the scope of this study, one could use the channel selection algorithm [69] to find geometric or anatomical patterns in the location of the most useful electrodes. These patterns could guide electrode placement for advanced prostheses used by persons with transradial amputation; however, the usefulness of the patterns would vary depending on the level of the amputation.

Our study did not directly measure or restrict compensatory movements. Future work would benefit from quantifying or restricting compensatory movements. For some tasks (e.g., pouring task), compensatory movements could make stable grip control more important to the user than the ability to simultaneously activate multiple degrees-of-freedom. One benefit of the bypass socket [68] used in this study is that the rotating wrist attachment generally keeps the prosthesis hanging below the user’s wrist, regardless of shoulder, elbow, or wrist orientation, thus reducing the ability of the user to make compensatory movements and hence indirectly requiring simultaneous movements. The clothespin inside-out task conducted in phase one is particularly difficult to complete with compensatory movements. As a **post-hoc** analysis of the clothespin task, we found that the mKF and MLP had more simultaneous movements than the CNN, suggesting that they may have required less compensation.

Reliable, simultaneous movements in position mode could provide the user with a prosthesis experience like that of the endogenous hand, in which simultaneous movements are natural. In velocity mode, a user may choose to complete multi-degree-of-freedom tasks sequentially, moving a single degree-of-freedom at a time. Sequential movements are far less natural and much slower than simultaneous movements if the simultaneous movements can be completed reliably. Our results suggest simultaneous movements were used for task completion; however, simultaneous movements alone do not mean the movements were intended. The mKF and the MLP had significantly more simultaneous movements than the CNN; however, reported user experiences and our informal observation suggest that the MLP may not have made the simultaneous movements reliably. The CNN, with less simultaneous movements, may have provided reliable grip control which benefited the task and was reflected in user experiences, although successful task performance would have required greater compensatory movements from the user.

Although common performance metrics were used, direct comparisons with other studies are difficult due to different experimental conditions (e.g., degrees-of-freedom, physical prosthesis). In [61], where a two-degree-of-freedom, continuous, linear regression-based control strategy was compared with a conventional control strategy, the participant required about 10 s per clothespin for the horizontal to vertical transfer with the regression-based approach, whereas the conventional strategy required about 14 s. In [49], the two-degree-of-freedom “proportional simultaneous” control strategy (most similar to what we have termed continuous control) required about 15 s to move a clothespin up and down (horizontal to vertical then vertical to horizontal), whereas the conventional strategy required about 29 s. Our median transfer times ranged from 7 s to 11 s to depending on the decoder, also two-degrees-of-freedom. Classifier approaches [20], [65], [66] on the clothespin relocation task have ranged from about 10 s to 30 s.

The fragile object task has been performed with various prostheses and fragile objects. Generally, the ratio between the break force and mass determines the task difficulty; a lower ratio implies greater difficulty. In the present study, non-amputee participants achieved a 63% median success rate. The break force in the present experiments was about 20 N, and the device mass was 615 g, resulting in a 0.03 N/g ratio. In a previous study from our group, an individual with an amputation achieved about 55% success with the mKF on a similar fragile egg task without sensory feedback [27]. In [86], [87], the success rates without sensory feedback were 38% and 45% with a fragile object that broke at 1.23 N and had a mass of 80 g, a ratio of 0.15 N/g, approximately five times the ratio of our study. In [88], the success rate was about 86% without sensory feedback, also using a different fragile object, which broke at 10.7 N and weighed 8 g, a 1.34 N/g ratio (approximately 45 times the ratio of our study). Transfer times, where reported, were similar among the aforementioned studies and the present study, ranging from 10 s to 13 s per transfer.

One goal of this study was to determine the best decoders and training paradigms for a portable take-home system. This study provides strong evidence from real-world functional tasks for using diverse training sets as input for a decoder and modifying the decode output with an LF. Selecting between linear and nonlinear decoders is less clear. Our findings indicate comparable performance between an mKF and CNN, where user preferences were slightly increased for the CNN. Clinical deployment, however, may favor the mKF, which trains faster and is less computationally expensive. Furthermore, in a take-home clinical trial where more training data may be collected over time, aggregation of these datasets may begin to favor deep learning approaches [67] like the CNN or MLP as technologies continue to improve. Overall, our results provide valuable information toward the clinical implementation of advanced control strategies for prosthesis users.

## Conclusions

This work compared myoelectric prosthetic control using activities of daily living with 12 continuous decoders in a two-phase study. In the first phase, comparing training paradigms and decoder smoothing, we found that training control algorithms with more complex movement patterns (i.e., simultaneously active digits and/or wrist) improves control accuracy and speed. Our results showed that nonlinear smoothing of the decoder output can improve accuracy although sometimes at a cost in speed. In the second phase, objective performance among the mKF, MLP, and CNN was similar for many tasks, but subjective preferences favored the CNN and mKF. Users’ subjective experiences are a key additional consideration.

This study was completed to select a decoder for a long-term take-home trial with implanted neuromyoelectric devices. The results demonstrate the importance of using rich training paradigms and nonlinear algorithm smoothing to improve continuous prosthetic control. The results also highlight the importance of user-focused subjective metrics in comparing decoders. Where performance differences could not effectively select a “best” decoder, user preferences added strong evidence toward selecting the mKF or CNN. Clinical considerations may favor the mKF as it is faster to train and computationally less expensive than the CNN. Subjective experiences may be even more important in long-term real-world environments and may influence voluntary prosthesis use. Overall, the results herein demonstrate the efficacy of continuous decoders for enabling intuitive prosthetic control of modern bionic prostheses.

## Data Availability

Available upon reasonable request.

## Abbreviations

EMG: electromyography
mKF: modified Kalman filter
MLP: multi-layer perceptron
CNN: convolutional neural network
sEMG: surface electromyography
TLX: Task Load Index

## Declarations

## Ethics approval and consent to participate

All participants took part in the experiments after giving their informed consent, as approved by the University of Utah IRB (ID 00098851).

## Consent for publication

Not applicable.

## Availability of data and materials

The datasets generated during and/or analyzed during the current study are available from the corresponding author on reasonable request.

## Competing interests

The author(s) declare(s) that they have no competing interests.

## Funding

This work was sponsored by the Hand Proprioception and Touch Interfaces (HAPTIX) program administered by the Biological Technologies Office (BTO) of the Defense Advanced Research Projects Agency (DARPA) through the Space and Naval Warfare Systems Center, Contract No. N66001-15-C-4017. Research reported in this publication was supported by the National Center for Advancing Translational Sciences of the National Institutes of Health (NIH) under Award Number UL1TR002538 and TL1TR002540. The content is solely the authors’ responsibility and does not necessarily represent the official views of their employers, DARPA, or the NIH.

## Authors’ contributions

MP designed the experiments, collected and analyzed the data, and wrote the manuscript. MB designed the experiments and collected the data. TH designed the experiments, collected and analyzed the data, and wrote the manuscript. JG optimized the mKF and implemented MLP and CNN algorithms. TD implemented mKF and MLP algorithms. CD provided experimental guidance and oversaw the study. GC oversaw all aspects of the study and writing. All authors contributed to revising the manuscript.

## Acknowledgments

We thank the participants in this study for their willingness to invest their time to improve advanced prostheses to benefit amputee populations.

## Additional Files

Video 1: Example videos of phase two tasks for each algorithm

## Notes

### Competing Interest Statement

The authors have declared no competing interest.

### Author Declarations

University of Utah IRB

### Summary of Updates

Added Table 1 including algorithm training times. Added new analysis on simultaneous movements.

## References

[1] Mobius Bionics LLC, “LUKE Arm System,” 2019. [Online]. Available: https://www.mobiusbionics.com/wp-content/uploads/2019/09/Mobius-Bionics-LUKE-Product-Spec-Sheet.pdf.

[2] Taska Prosthetics, “TASKA Hand,“ 2020. https://www.taskaprosthetics.com/the-taska (accessed Jul. 08, 2020).

[3] Otto Bock HealthCare LP, “bebionic hand User Guide,” 2018. [Online]. Available: https://www.ottobock.ca/media/local-media/prosthetics/upper-limb/files/15667-bebionic-user-guide.pdf.

[4] Otto Bock HealthCare LP, “Michelangelo Hand Brochure,” 2017. [Online]. Available: http://www.ottobockus.com/media/local-media/prosthetics/upper-limb/michelangelo/files/michelangelo-brochure.pdf.

[5] Motion Control Division of Fillauer, “Utah Arm U3 and U3+ User Guide,” 2020.

[6] Open Bionics LLC, “Hero Arm -User Guide,” 2020. https://openbionics.com/hero-arm-user-guide/ (accessed Jul. 08, 2020).

[7] Össur Touch Solutions, “i-Limb Quantum Titanium,” 2020.

[8] Össur Touch Solutions, “i-Limb Access,” 2020.

[9] S. M. Engdahl, B. P. Christie, B. Kelly, A. Davis, C. A. Chestek, and D. H. Gates, “Surveying the interest of individuals with upper limb loss in novel prosthetic control techniques,” J. Neuroeng. Rehabil., vol. 12, no. 1, pp. 1–11, 2015, doi: 10.1186/s12984-015-0044-2.

[10] E. Biddiss and T. Chau, “Upper limb prosthesis use and abandonment: A survey of the last 25 years,” Prosthet. Orthot. Int., vol. 31, no. 3, pp. 236–257, 2007, doi: 10.1080/03093640600994581.

[11] Coapt LLC, “Complete Control System Gen2 Handbook.” 2020.

[12] Otto Bock HealthCare LP, “Myo Plus pattern recognition,” 2020..

[13] M. Asghari Oskoei and H. Hu, “Myoelectric control systems-A survey,” Biomed. Signal Process. Control, vol. 2, no. 4, pp. 275–294, 2007, doi: 10.1016/j.bspc.2007.07.009.

[14] Touchbionics, “I-limb quantum,” 2018. http://www.touchbionics.com/products/active-prostheses/i-limb-quantum (accessed Mar. 07, 2019).

[15] E. D’Anna et al., “A closed-loop hand prosthesis with simultaneous intraneural tactile and position feedback,” Sci. Robot., vol. 4, no. 27, 2019, doi: 10.1126/scirobotics.aau8892.

[16] T. A. Kuiken et al., “Targeted muscle reinnervation for real-time myoelectric control of multifunction artificial arms,” JAMA - J. Am. Med. Assoc., vol. 301, no. 6, pp. 619–628, 2009, doi: 10.1001/jama.2009.116.

[17] A. Waris, I. K. Niazi, M. Jamil, K. Englehart, W. Jensen, and E. N. Kamavuako, “Multiday Evaluation of Techniques for EMG-Based Classification of Hand Motions,” IEEE J. Biomed. Heal. Informatics, vol. 23, no. 4, pp. 1526–1534, 2019, doi: 10.1109/JBHI.2018.2864335.

[18] A. J. Young, L. H. Smith, E. J. Rouse, and L. J. Hargrove, “Classification of simultaneous movements using surface EMG pattern recognition,” IEEE Trans. Biomed. Eng., vol. 60, no. 5, pp. 1250–1258, 2013, doi: 10.1109/TBME.2012.2232293.

[19] L. J. Hargrove, G. Li, K. B. Englehart, and B. S. Hudgins, “Principal components analysis preprocessing for improved classification accuracies in pattern-recognition-based myoelectric control,” IEEE Trans. Biomed. Eng., vol. 56, no. 5, pp. 1407–1414, 2009, doi: 10.1109/TBME.2008.2008171.

[20] L. J. Hargrove, L. A. Miller, K. Turner, and T. A. Kuiken, “Myoelectric Pattern Recognition Outperforms Direct Control for Transhumeral Amputees with Targeted Muscle Reinnervation: A Randomized Clinical Trial,” Sci. Rep., vol. 7, no. 1, pp. 1–9, 2017, doi: 10.1038/s41598-017-14386-w.

[21] E. D’Anna et al., “A somatotopic bidirectional hand prosthesis with transcutaneous electrical nerve stimulation based sensory feedback,” Sci. Rep., vol. 7, no. 1, pp. 1–15, Dec. 2017, doi: 10.1038/s41598-017-11306-w.

[22] M. Ortiz-Catalan, B. Håkansson, and R. Brånemark, “An osseointegrated human-machine gateway for long term sensory feedback and control of artificial limbs,” Sci. Transl. Med., vol. 6, no. 257, pp. 1–9, 2014.

[23] E. Scheme and K. Englehart, “Training strategies for mitigating the effect of proportional control on classification in pattern recognition-based myoelectric control,” J. Prosthetics Orthot., vol. 25, no. 2, pp. 76–83, 2013, doi: 10.1097/JPO.0b013e318289950b.

[24] N. Jiang, K. B. Englehart, and P. A. Parker, “Extracting simultaneous and proportional neural control information for multiple-dof prostheses from the surface electromyographic signal,” IEEE Trans. Biomed. Eng., vol. 56, no. 4, pp. 1070–1080, 2009, doi: 10.1109/TBME.2008.2007967.

[25] W. Wu, Y. Gao, E. Bienenstock, J. P. Donoghue, and M. J. Black, “Bayesian population decoding of motor cortical activity using a Kalman filter,” Neural Comput., vol. 18, no. 1, pp. 80–118, 2006, doi: 10.1162/089976606774841585.

[26] P. P. Vu et al., “A regenerative peripheral nerve interface allows real-time control of an artificial hand in upper limb amputees,” Sci. Transl. Med., vol. 12, no. 533, pp. 1–12, 2020, doi: 10.1126/scitranslmed.aay2857.

[27] J. A. George et al., “Biomimetic sensory feedback through peripheral nerve stimulation improves dexterous use of a bionic hand,” Sci. Robot., vol. 4, no. 32, p. eaax2352. Jul. 2019, doi: 10.1126/scirobotics.aax2352.

[28] A. Ameri, E. N. Kamavuako, E. J. Scheme, K. B. Englehart, and P. A. Parker, “Support vector regression for improved real-time, simultaneous myoelectric control,” IEEE Trans. Neural Syst. Rehabil. Eng., vol. 22, no. 6, pp. 1198–1209, 2014, doi: 10.1109/TNSRE.2014.2323576.

[29] H. Dantas, D. J. Warren, S. M. Wendelken, T. S. Davis, G. A. Clark, and V. J. Mathews, “Deep Learning Movement Intent Decoders Trained with Dataset Aggregation for Prosthetic Limb Control,” IEEE Trans. Biomed. Eng., vol. 66, no. 11, pp. 3192–3203, 2019, doi: 10.1109/TBME.2019.2901882.

[30] J. A. George, M. R. Brinton, C. C. Duncan, D. T. Hutchinson, and G. A. Clark, “Improved Training Paradigms and Motor-decode Algorithms: Results from Intact Individuals and a Recent Transradial Amputee with Prior Complex Regional Pain Syndrome,” in 40th International Engineering in Medicine and Biology Conference, 2018, vol. 2018, doi: 10.1109/EMBC.2018.8513342.

[31] A. Ameri, M. A. Akhaee, E. Scheme, and K. Englehart, “Regression convolutional neural network for improved simultaneous EMG control,” J. Neural Eng., 2019, doi: 10.1088/1741-2552/ab0e2e.

[32] A. A. Adewuyi, L. J. Hargrove, and T. A. Kuiken, “Resolving the effect of wrist position on myoelectric pattern recognition control,” J. Neuroeng. Rehabil., vol. 14, no. 1, 2017, doi: 10.1186/s12984-017-0246-x.

[33] T. C. Hansen, H. Dantas, J. A. George, G. A. Clark, D. J. Warren, and V. J. Mathews, “Shared Controllers Improve Control and Performance of Upper-Limb Prostheses,” 2019, doi: 10.13140/RG.2.2.36537.72805.

[34] V. Gilja et al., “A high-performance neural prosthesis enabled by control algorithm design,” Nature Neuroscience, vol. 15, no. 12. pp. 1752–1757, 2012, doi: 10.1038/nn.3265.

[35] J. A. George, T. S. Davis, M. R. Brinton, and G. A. Clark, “Intuitive neuromyoelectric control of a dexterous bionic arm using a modified Kalman filter,” J. Neurosci. Methods, vol. 330, no. April 2019, p. 108462, 2020, doi: 10.1016/j.jneumeth.2019.108462.

[36] M. Ortiz-Catalan, B. Håkansson, and R. Brånemark, “Real-time and simultaneous control of artificial limbs based on pattern recognition algorithms,” IEEE Trans. Neural Syst. Rehabil. Eng., vol. 22, no. 4, pp. 756–764, 2014, doi: 10.1109/TNSRE.2014.2305097.

[37] L. J. Hargrove, K. Englehart, and B. Hudgins, “A comparison of surface and intramuscular myoelectric signal classification,” IEEE Trans. Biomed. Eng., vol. 54, no. 5, pp. 847–853, 2007, doi: 10.1109/TBME.2006.889192.

[38] A. Ameri, E. N. Kamavuako, E. J. Scheme, K. B. Englehart, and P. A. Parker, “Real-time, simultaneous myoelectric control using visual target-based training paradigm,” Biomed. Signal Process. Control, vol. 13, no. 1, pp. 8–14, 2014, doi: 10.1016/j.bspc.2014.03.006.

[39] E. N. Kamavuako, K. B. Englehart, W. Jensen, and D. Farina, “Simultaneous and proportional force estimation in multiple degrees of freedom from intramuscular EMG,” IEEE Trans. Biomed. Eng., vol. 59, no. 7, pp. 1804–1807, 2012, doi: 10.1109/TBME.2012.2197210.

[40] J. M. Hahne et al., “Linear and Nonlinear Regression Techniques for Simultaneous and Proportional Myoelectric Control,” IEEE Trans. Neural Syst. Rehabil. Eng., vol. 22, no. 2, pp. 269–279, Mar. 2014, doi: 10.1109/TNSRE.2014.2305520.

[41] A. Ameri, M. A. Akhaee, E. Scheme, and K. Englehart, “Real-time, simultaneous myoelectric control using a convolutional neural network,” PLoS One, vol. 13, no. 9, pp. 1–13, 2018, doi: 10.1371/journal.pone.0203835.

[42] M. Zia ur Rehman et al., “Multiday EMG-Based classification of hand motions with deep learning techniques,” Sensors (Switzerland), vol. 18, no. 8, pp. 1–16, 2018, doi: 10.3390/s18082497.

[43] A. Fougner, E. Scheme, A. D. C. Chan, K. Englehart, and Ø. Stavdahl, “Resolving the limb position effect in myoelectric pattern recognition,” IEEE Trans. Neural Syst. Rehabil. Eng., vol. 19, no. 6, pp. 644–651, 2011, doi: 10.1109/TNSRE.2011.2163529.

[44] Y. Teh and L. J. Hargrove, “Understanding limb position and external load effects on real-time pattern recognition control in amputees,” IEEE Trans. Neural Syst. Rehabil. Eng., vol. 28, no. 7, pp. 1–1, 2020, doi: 10.1109/tnsre.2020.2991643.

[45] A. Radmand, E. Scheme, and K. Englehart, “On the suitability of integrating accelerometry data with electromyography signals for resolving the effect of changes in limb position during dynamic limb movement,” J. Prosthetics Orthot., vol. 26, no. 4, pp. 185–193, 2014, doi: 10.1097/JPO.0000000000000041.

[46] D. Young et al., “Closed-loop cortical control of virtual reach and posture using Cartesian and joint velocity commands,” J. Neural Eng., vol. 16, no. 2, 2019, doi: 10.1088/1741-2552/aaf606.

[47] J. E. Downey et al., “Blending of brain-machine interface and vision-guided autonomous robotics improves neuroprosthetic arm performance during grasping,” J. Neuroeng. Rehabil., vol. 13, no. 1, pp. 1–12, 2016, doi: 10.1186/s12984-016-0134-9.

[48] J. Nieveen, M. R. Brinton, D. J. Warren, and V. J. Mathews, “A Nonlinear Latching Filter to Remove Jitter From Movement Estimates for Prostheses,” IEEE Trans. Neural Syst. Rehabil. Eng., 2020, doi: 10.1109/TNSRE.2020.3038706.

[49] A. L. Fougner, Ø. Stavdahl, and P. J. Kyberd, “System training and assessment in simultaneous proportional myoelectric prosthesis control,” J. Neuroeng. Rehabil., vol. 11, no. 1, pp. 1–13, 2014, doi: 10.1186/1743-0003-11-75.

[50] M. Ortiz-Catalan, F. Rouhani, R. Branemark, and B. Hakansson, “Offline accuracy: A potentially misleading metric in myoelectric pattern recognition for prosthetic control,” Proc. Annu. Int. Conf. IEEE Eng. Med. Biol. Soc. EMBS, vol. 2015-Novem, pp. 1140–1143, 2015, doi: 10.1109/EMBC.2015.7318567.

[51] E. Scheme and K. Englehart, “On the robustness of EMG features for pattern recognition based myoelectric control; A multi-dataset comparison,” 2014 36th Annu. Int. Conf. IEEE Eng. Med. Biol. Soc. EMBC 2014, pp. 650–653, 2014, doi: 10.1109/EMBC.2014.6943675.

[52] M. Cracchiolo et al., “Decoding of grasping tasks from intraneural recordings in trans-radial amputee,” J. Neural Eng., vol. 17, no. 2, p. 026034, 2020, doi: 10.1088/1741-2552/ab8277.

[53] N. Jiang, I. Vujaklija, H. Rehbaum, B. Graimann, and D. Farina, “Is accurate mapping of EMG signals on kinematics needed for precise online myoelectric control?,” IEEE Trans. Neural Syst. Rehabil. Eng., vol. 22, no. 3, pp. 549–558, 2014, doi: 10.1109/TNSRE.2013.2287383.

[54] P. P. Vu et al., “Closed-Loop Continuous Hand Control via Chronic Recording of Regenerative Peripheral Nerve Interfaces,” IEEE Trans. Neural Syst. Rehabil. Eng., vol. 26, no. 2, pp. 515–526, Feb. 2018, doi: 10.1109/TNSRE.2017.2772961.

[55] N. Jiang, T. Lorrain, and D. Farina, “A state-based, proportional myoelectric control method: online validation and comparison with the clinical state-of-the-art,” J. Neuroeng. Rehabil., vol. 11, no. 1, p. 110, 2014, doi: 10.1186/1743-0003-11-110.

[56] J. A. Birdwell, L. J. Hargrove, R. F. F. Weir, and T. A. Kuiken, “Extrinsic Finger and Thumb Muscles Command a Virtual Hand to Allow Individual Finger and Grasp Control,” IEEE Trans. Biomed. Eng., vol. 62, no. 1, pp. 218–226, 2014, doi: 10.1016/j.physbeh.2017.03.040.

[57] L. H. Smith, T. A. Kuiken, and L. J. Hargrove, “Real-time simultaneous and proportional myoelectric control using intramuscular EMG,” J. Neural Eng., vol. 11, no. 6, 2014, doi: 10.1088/1741-2560/11/6/066013.

[58] I. Vujaklija et al., “Translating research on myoelectric control into clinics-are the performance assessment methods adequate?,” Front. Neurorobot., vol. 11, no. FEB, 2017, doi: 10.3389/fnbot.2017.00007.

[59] J. L. Betthauser et al., “Limb Position Tolerant Pattern Recognition for Myoelectric Prosthesis Control with Adaptive Sparse Representations from Extreme Learning,” IEEE Trans. Biomed. Eng., vol. 65, no. 4, pp. 770–778, 2018, doi: 10.1109/TBME.2017.2719400.

[60] L. Hargrove, L. Miller, K. Turner, and T. Kuiken, “Control within a virtual environment is correlated to functional outcomes when using a physical prosthesis,” J. Neuroeng. Rehabil., vol. 15, no. Suppl 1, 2018, doi: 10.1186/s12984-018-0402-y.

[61] J. M. Hahne, M. A. Wilke, M. Koppe, D. Farina, and A. F. Schilling, “Longitudinal Case Study of Regression-Based Hand Prosthesis Control in Daily Life,” Front. Neurosci., vol. 14, no. June, pp. 1–8, 2020, doi: 10.3389/fnins.2020.00600.

[62] J. M. Hahne, M. A. Schweisfurth, M. Koppe, and D. Farina, “Simultaneous control of multiple functions of bionic hand prostheses: Performance and robustness in end users,” Sci. Robot., vol. 3, no. 19, p. eaat3630, 2018, doi: 10.1126/scirobotics.aat3630.

[63] S. Amsuess, P. Goebel, B. Graimann, and D. Farina, “A multi-class proportional myocontrol algorithm for upper limb prosthesis control: Validation in real-life scenarios on amputees,” IEEE Trans. Neural Syst. Rehabil. Eng., vol. 23, no. 5, pp. 827–836, 2015, doi: 10.1109/TNSRE.2014.2361478.

[64] T. A. Kuiken et al., “Targeted reinnervation for enhanced prosthetic arm function in a woman with a proximal amputation: a case study,” Lancet, vol. 369, no. 9559, pp. 371–380, 2007, doi: 10.1016/S0140-6736(07)60193-7.

[65] T. A. Kuiken, L. A. Miller, K. Turner, and L. J. Hargrove, “A Comparison of Pattern Recognition Control and Direct Control of a Multiple Degree-of-Freedom Transradial Prosthesis,” IEEE J. Transl. Eng. Heal. Med., vol. 4, 2016, doi: 10.1109/JTEHM.2016.2616123.

[66] G. K. Patel, C. Castellini, J. M. Hahne, D. Farina, and S. Dosen, “A Classification Method for Myoelectric Control of Hand Prostheses Inspired by Muscle Coordination,” IEEE Trans. Neural Syst. Rehabil. Eng., vol. 26, no. 9, pp. 1745–1755, 2018, doi: 10.1109/TNSRE.2018.2861774.

[67] J. A. George, A. Neibling, M. D. Paskett, and G. A. Clark, “Inexpensive surface electromyography sleeve with consistent electrode placement enables dexterous and stable prosthetic control through deep learning,” 41st Int. Eng. Med. Biol. Conf., vol. 2020, 2020, [Online]. Available: http://arxiv.org/abs/2003.00070.

[68] M. D. Paskett et al., “A Modular Transradial Bypass Socket for Surface Myoelectric Prosthetic Control in Non-Amputees,” IEEE Trans. Neural Syst. Rehabil. Eng., vol. 27, no. 10, pp. 2070–2076, 2019, doi: 10.1109/TNSRE.2019.2941109.

[69] J. Nieveen, D. Warren, S. Wendelken, T. Davis, D. Kluger, and D. Page, “Channel Selection of Neural And Electromyographic Signals for Decoding of Motor Intent,” in Myoelectric Controls Conference, 2017, p. 720.

[70] A. Hussaini and P. Kyberd, “Refined clothespin relocation test and assessment of motion,” Prosthet. Orthot. Int., vol. 41, no. 3, pp. 294–302, 2017, doi: 10.1177/0309364616660250.

[71] N. R. Olsen et al., “An Adaptable Prosthetic Wrist Reduces Subjective Workload,” bioRxiv, 2019, doi: 10.1101/808634.

[72] S. G. Hart and L. E. Staveland, “Development of NASA-TLX (Task Load Index): Results of Empirical and Theoretical Research,” Adv. Psychol., 1988, doi: 10.1016/S0166-4115(08)62386-9.

[73] J. Peat and B. Barton, Medical Statistics: A Guide to Data Analysis and Critical Appraisal. 2008.

[74] J. D. Gibbons and S. Chakraborti, “Nonparametric Statistical Inference,” in International Encyclopedia of Statistical Science, 2011.

[75] R. A. Armstrong, “When to use the Bonferroni correction,” Ophthalmic Physiol. Opt., vol. 34, no. 5, pp. 502–508, 2014, doi: 10.1111/opo.12131.

[76] R. V. Hogg and J. Ledolter, Engineering Statistics. New York: MacMillan, 1987.

[77] Y. Hochberg and A. C. Tamhane, Multiple Comparison Procedures, 1st ed. John Wiley & Sons, Inc., 1987.

[78] S. G. Meek, S. C. Jacobsen, and P. P. Goulding, “Extended physiologic taction: Design and evaluation of a proportional force feedback system,” J. Rehabil. Res. Dev., vol. 26, no. 3, pp. 53–62, 1989.

[79] E. D. Engeberg and S. Meek, “Improved grasp force sensitivity for prosthetic hands through force-derivative feedback,” IEEE Trans. Biomed. Eng., vol. 55, no. 2, pp. 817–821, 2008, doi: 10.1109/TBME.2007.912675.

[80] W. H. Kruskal and W. A. Wallis, “Use of Ranks in One-Criterion Variance Analysis,” J. Am. Stat. Assoc., 1952, doi: 10.1080/01621459.1952.10483441.

[81] Z. Šidák, “Rectangular Confidence Regions for the Means of Multivariate Normal Distributions,” J. Am. Stat. Assoc., 1967, doi: 10.1080/01621459.1967.10482935.

[82] S. Muceli and D. Farina, “Simultaneous and proportional estimation of hand kinematics from EMG during mirrored movements at multiple degrees-of-freedom,” IEEE Trans. Neural Syst. Rehabil. Eng., vol. 20, no. 3, pp. 371–378, 2012, doi: 10.1109/TNSRE.2011.2178039.

[83] J. A. George, T. N. Tully, P. Colgan, and G. Clark, “Bilaterally Mirrored Movements Improve the Accuracy and Precision of Training Data for Supervised Learning of Neural or Myoelectric Prosthetic Control,” 2020, doi: 10.13140/RG.2.2.34651.59689.

[84] F. Cini, V. Ortenzi, P. Corke, and M. Controzzi, “On the choice of grasp type and location when handing over an object,” Sci. Robot., vol. 4, no. 27, pp. 1–14, 2019, doi: 10.1126/scirobotics.aau9757.

[85] S. Sundaram, P. Kellnhofer, Y. Li, J.-Y. Zhu, A. Torralba, and W. Matusik, “Learning the signatures of the human grasp using a scalable tactile glove,” Nature, vol. 569, no. 7758, pp. 698–702, 2019, doi: 10.1038/s41586-019-1234-z.

[86] G. Valle et al., “Biomimetic Intraneural Sensory Feedback Enhances Sensation Naturalness, Tactile Sensitivity, and Manual Dexterity in a Bidirectional Prosthesis,” Neuron, vol. 100, no. 1, pp. 37-45.e7, Oct. 2018, doi: 10.1016/j.neuron.2018.08.033.

[87] F. M. Petrini et al., “Six-Month Assessment of a Hand Prosthesis with Intraneural Tactile Feedback,” Ann. Neurol., vol. 85, no. 1, pp. 137–154, 2019, doi: 10.1002/ana.25384.

[88] F. Clemente, M. D’Alonzo, M. Controzzi, B. B. Edin, and C. Cipriani, “Non-Invasive, Temporally Discrete Feedback of Object Contact and Release Improves Grasp Control of Closed-Loop Myoelectric Transradial Prostheses,” IEEE Trans. Neural Syst. Rehabil. Eng., vol. 24, no. 12, pp. 1314–1322, 2016, doi: 10.1109/TNSRE.2015.2500586.

